# Gray matter microstructure from in-vivo diffusion MRI reflects post-mortem neuropathology severity and clinical progression of Alzheimer’s disease

**DOI:** 10.1101/2025.05.30.25328630

**Authors:** Zaki Alasmar, Cécilia Tremblay, Roqaie Moqadam, Geidy E Serrano, Thomas G Beach, Alizera Atri, Yi Su, the Alzheimer’s Disease Neuroimaging Initiative, Yashar Zeighami, Mahsa Dadar

**Author notes:** These authors contributed equally to this work (Zaki Alasmar, Cécilia Tremblay). co-senior authors (Yashar Zeighami, Mahsa Dadar). Data used in preparation for this article were obtained from the Alzheimer’s Disease Neuroimaging Initiative (ADNI) database (adni.loni.usc.edu). As such, the investigators within the ADNI contributed to the design and implementation of ADNI and/or provided data but did not participate in the analysis or writing of this report. A complete listing of ADNI investigators can be found at: http://adni.loni.usc.edu/wpontent/uploads/how_to_apply/ADNI_Acknowledgement_List.pdf. **CORRESPONDENCE:** Mahsa Dadar, Cerebral Imaging Centre, Douglas Mental Health University Institute, Montréal, Québec, Canada, H4H 1R3 Yashar Zeighami, Cerebral Imaging Centre, Douglas Mental Health University Institute, Montréal, Québec, Canada, H4H 1R3.

## Abstract

**INTRODUCTION:** Diffusion-weighted imaging derived mean diffusivity (MD) correlates with Alzheimer’s disease biomarkers, yet its neuropathological correlates remain unclear.

**METHODS:** Diffusion-weighted imaging, postmortem neuropathology, and cognitive performance data were obtained from the National Alzheimer’s Coordinating Center (N=97), Alzheimer’s Disease Neuroimaging Initiative (N=21), and Arizona Study of Aging and Neurodegenerative Disorders (N=15). We examined MD associations with neuropathology, cognitive decline, and expression profiles of AD-implicated genes.

**RESULTS:** Results revealed two latent variables—one linked to amyloid/tau, the other to vascular pathology—explaining 70% and 16% of MD-pathology covariance, respectively. Higher MD correlated with worse cognitive performance, both cross-sectionally and up to 14 years prior to death. MD was regionally associated with Thal phase, neuritic plaque density, Braak stage (temporal/limbic), and infarcts (thalamus), and reflected gene expression patterns related to AD.

**DISCUSSION:** In vivo MD captures distinct AD-related pathologies across brain regions and relates to cognitive trajectories and gene expression.

## 1. BACKGROUND

Neuropathological changes in Alzheimer’s Disease (AD) begin decades before symptom onset [1]. Due to the heterogeneous nature of AD-related changes in their early phases, methods that can detect these changes are needed to identify individuals at risk of developing AD. This enables early interventions that have a potential to delay or prevent cognitive decline [2]. Despite great developmental advances of biofluid and positron emission tomography (PET) biomarkers, there is still a critical need for the sensitive, less invasive, and widely applicable in-vivo biomarkers that can estimate AD pathology burden and predict clinical progression, cognitive decline and possibly serve as suitable outcome measures for clinical trials [3].

There is increasing evidence that early AD neuropathology, including accumulation of amyloid β plaques, tau neurofibrillary tangles, and neuroinflammation cause microstructural changes that may be detected by Magnetic Resonance Imaging (MRI) [4,5]. In fact, Diffusion-Weighted Imaging (DWI) methods have been more recently used to assess cortical microstructural grey matter changes across neurodegenerative diseases [6–8]. The diffusivity changes in grey matter, as measured by DWI-derived mean diffusivity (MD), have been found to be sensitive to microstructural changes, and can be detected earlier than macrostructural neurodegeneration and associated volumetric changes [1,9]. In clinically manifested AD and mild cognitive impairment (MCI), increased MD values were reported in hippocampal regions as well as other cortical regions associated with cognitive decline [5,7,10]. DWI measures were further found to predict cognitive decline and were associated with cerebrospinal fluid (CSF) based AD biomarkers [5,11,12]. Microstructural cortical changes were also shown to correlate with AD markers as measured by amyloid β and tau PET [12,13]. Further, widespread increases in cortical MD were observed over time in temporal and parietal cortical regions in amyloid β positive cases, indicating that MD can serve as an sensitive marker of early microstructural change in AD [14,15].

While these findings have been promising, there are limitations to PET and fluid biomarkers with regards to quantifying neuropathology, and their sensitivity and accuracy may vary depending on disease stages [11,16–19]. Furthermore, fluid biomarkers do not provide any information on the spatial location of the pathologies, while current PET tracers are not able to distinguish diffuse plaques from neuritic plaques [20]. These limitations prevent accurate localization of neuropathology in the brain and longitudinal tracking of the impact of different types of neuropathologies on cognitive dysfunction and disease progression [21]. Hence, postmortem pathological validation is still critically needed to examine DWI measures as markers of neuropathology burden in AD-related regions and to better understand the underlying processes affecting them. Such clinicopathological studies are sparse and only one study has demonstrated a link between cortical DWI and AD neuropathologic changes in a cohort of 43 autopsy cases [22]. Current studies do not account for the presence of common co-pathologies, especially vascular pathology, that is frequently observed at autopsy [23]. Considering these factors would improve the ability to discriminate between different pathologies and/or the presence of multiple comorbidities in AD.

In this clinicopathological study, we aimed to assess the link between microstructural changes, as measured by MD, and postmortem amyloid, tau, and vascular burden and distribution in pathologically characterized aging individuals along the AD neuropathological continuum. To do so, we used data from three sources including the National Alzheimer’s Coordinating Center (NACC), Alzheimer’s Disease Neuroimaging Initiative (ADNI), and the Arizona Study of Aging and Neurodegenerative Disorders (AZSAND) and its Brain and Body Donation Program (BBDP) [24–28]. We hypothesized that MD would be associated with regional amyloid and tau burden as measured by neuropathological stagings including Thal amyloid phase, neuritic and diffuse plaque density, and Braak neurofibrillary tangle staging as well as vascular pathology. We further hypothesized it will reflect longitudinal ante-mortem clinical measures of cognitive functioning. Lastly, we hypothesized that our derived MD pattern is associated with expression levels of genes implicated in AD pathology.

## 2. METHODS

### 2.1 Overview of approach and study population

To assess the link between MD and neuropathology, all participants with available DWI and neuropathological assessments after death were selected from three different sources. We aggregated participants’ data from the NACC (September 2024 data freeze; https://naccdata.org/; N=109) [25,28], ADNI (https://adni.loni.usc.edu/; N=22) a longitudinal multicenter observational study aiming to validate biomarkers of AD [26,27] as well as additional participants from the Arizona Study of Aging and Neurodegenerative Disorders (AZSAND) and its Brain and Body Donation Program (BBDP), a longitudinal clinicopathological study at Banner Sun Health Research Institute (BSHRI) (http://www.brainandbodydonationprogram.org/; N=16) [24]. Ethics approval and consent was obtained from the participants by the local institutional review board at the National Institute on Aging Alzheimer’s Disease Centers (ADRCs; which contributed the data to NACC), and for the ADNI and AZSAND participants. In addition to MRI data, all participants underwent clinical assessments and had Clinical Dementia Rating (CDR) information available. In total, we had 147 participants whose mean time between MRI and death was 5.14 ± 3.10 years (range = [0-13 years]).

### 2.2 Neuropathological assessments

Autopsies and neuropathological assessments were performed by the ADRCs contributing data to NACC, at the central neuropathology laboratory of the ADNI, or at BISHRI for AZSAND/BBDP cases. Arizona ADRC subjects are enrolled and autopsied in AZSAND/BBDP. The NACC Neuropathology Data Form was used by all centers which ensured consistency across datasets [25,28].

Neuropathology assessments included the rating scales used to classify core AD neuropathological hallmarks as the AD neuropathologic change according to the National Institute on Aging and Alzheimer’s Association (NIA-AA) guidelines [29,30]. These includes the distribution of senile plaques classified by Thal amyloid phase (ranging from 0 to 5; NACC variable NPTHAL) [31], the neurofibrillary tau tangle distribution classified by Braak stage (ranging from 0 to 6; NACC variable NPBRAAK/NACCBRAA) [32,33], and the density of neuritic plaques according to the Consortium to Establish a Registry for AD (CERAD) (ranging from 0 to 3; NACC variable NPNEUR/NACCNEUR) [34]. Additionally, diffuse plaque density classified analogously to CERAD neuritic plaque (ranging from 0 to 3; NACC variable NPDIFF/NACCDIFF), cerebral amyloid angiopathy (ranging from 0 to 3; NACC variable NPAMY), white matter rarefaction (ranging from 0 to 3; NPWMR), presence or absence of old gross or lacunar infarcts (NACC variable NPINF) or old microscopic infarcts (NACC variable NPOLD) and presence of vascular pathology (ischemic, hemorrhagic, or vascular pathology; (NACC variable NACCVASC) were also investigated.

### 2.3 MRI acquisition protocols

Structural T1w and DWI images were obtained for the 3 datasets. For NACC, all T1w scans were acquired with a gradient echo protocol, with 1 mm^3^ isotropic or 1×1×1.2 mm^3^ voxel size. DWI scans were all acquired with axial slices, with voxel size of either 2×2×3 mm^3^ or 1×1×2.5 mm^3^. b-values and the number of acquired directions varied across participants and included b=1000s/mm^2^ at 60 unique directions, b=1300s/mm^2^ with 40 unique directions, and some scans were conducted at higher gradient strength of b=3000s/mm^2^ with 64 unique directions. All DWI images were preceded by 2 or more non-diffusion weighted scans (B0 scans). For ADNI, all T1w images were acquired with TE=3ms, TR=7.3ms, 1 mm^3^ isotropic resolution. DWI were acquired with axial slices, with 1.4×1.4×2.7 mm^3^ voxel resolution and b-value of 1000 with 41 unique directions. All diffusion-weighted volumes were preceded by at least 2 B0 scans. Lastly, for AZSAND data, T1w scans were acquired with 1.2×1×1 mm^3^ voxel sizes. DWI volumes were acquired with axial slices with the following parameters: TE=57ms, TR=9s, 1.4×1.4×2.7 mm^3^ voxel size. All DWI scans were acquired with b=1000s/mm^2^ with 41 unique directions, which were preceded with 5 B0 volumes. A subsequent reverse-phase B0 scan with the same acquisition parameters was also acquired.

### 2.4 MRI preprocessing

All T1w scans were preprocessed using the same pipeline, which included denoising, intensity inhomogeneity correction, and intensity normalization using the minc-Toolkit v2 (https://bic-mni.github.io/; [35,36]). The preprocessed T1w images were then registered to MNI-ICBM152 template, using affine linear transformations followed by nonlinear deformations using Advanced Normalization Tools (ANTs; [37,38]. The transformations and warps fields were then applied to the Allen Human Brain Atlas [39] to map it to each participant’s native T1w image.

For DWI analysis, we used the Mrtrix3 and FSL package suits to preprocess the diffusion volumes and extract MD [40–42]. All DWI volumes first underwent Marchenko-Pastur PCA denoising, ringing artifact removal, susceptibility and intervolume motion correction [42,43]. To correct for phase shift artifacts in the NACC and ADNI datasets, we created a reverse-phase encoding non-diffusion weighted volume (i.e. reverse phase B0) via the Synthb0-Disco tool given that they lack an acquired no reverse-phase B0 [44,45]. This was done via accurate rigid alignment of the phase B0 with the T1w scan, which was then used to generate an undistorted B0 image via a pre-trained generative network. Given that the BBDP data were acquired with a reverse phase B0, we used it to correct for susceptibility artifacts rather than synthesizing a B0. We then used ANTs N4 to perform bias correction of the volumes, and compute MD at each voxel from the fitted diffusion tensor. The Allen atlas was then aligned to each participant’s MD map via the ANTs linear DWI-T1w co-registration transforms, and median MD values were extracted per region.

### 2.5 Quality control and imaging data harmonization

Atlas registration and the quality of the tensor metrics were visually inspected by two raters (ZA and MD) blind to participants’ diagnoses and neuropathology information. Six cases from NACC, one case from BBDP, and one case from ADNI failed registration and normalization steps and their data were excluded. Median MD per Allen Atlas region was aggregated for the remaining participants, and an iterative approach was used to impute missing values. We used multiple imputation by chained equations to impute missing regions’ MD values, based on the expected fit from all other regions. In this approach, median MD in each region is regressed against all other regions, and missing values are imputed with the expected MD based on the least squares solution [46–48]. We then harmonized regional MD across different acquisition protocols using neuroCombat (DWI specific; [49]). Neurocombat is a data harmonization tool developed specifically for neuroimaging, which was adapted from gene expression analysis [50]. It corrects for variations across data sources without removing effects of interest. Here, we followed a parametric empirical Bayes procedure to harmonize the means and variances of MD across all participants, while including the participants’ sex as a biological factor to conserve. In total, 6 NACC participants failed this step and were excluded from the rest of the analyses. After preprocessing, data quality check, imputation, and harmonization, the final sample size included 133 participants (97 NACC, 21 ADNI, and 15 BBDP).

### 2.5 Statistical Analysis

#### 2.5.1 Multivariate association between md and neuropathology

In order to assess the association between neuropathology markers and MD patterns across all brain regions, we conducted a multivariate regression analysis combining all the neuropathology markers and regions in one model. We used partial-least squares singular value decomposition (PLS-SVD) to examine the association between neuropathology and regional MD [51,52]. This procedure computes the covariance between MD of brain regions and neuropathologies, and extracts latent variables (LVs) of the underlying brain-neuropathology associations. Each latent variable is therefore a composite of regional MD scores (i.e. brain-wide MD) and combined neuropathologies and captures a specific proportion of these associations. In this analysis, we included age at death, sex, and time between DWI acquisition and death as covariates of no interest. We performed 1000 permutations on the latent variables to identify the null distribution, and 1000 bootstraps to model the 95% confidence intervals on the contribution of each region/neuropathological marker to the latent variables. A threshold of p < 0.05 after permutation was used to determine statistical significance.

#### 2.5.2 Univariate associations between MD and neuropathology

We further investigated the relationship between regional MD and each neuropathological marker using a univariate approach. Pearson’s correlations coefficients were computed between regional MD values and neuropathological markers, including Thal phase, Braak stage, neuritic plaque density, diffuse plaque density, cerebral amyloid angiopathy severity, and cerebral white matter rarefaction. For binary outcomes including the presence of infarcts (microinfarcts and/or gross, including lacunar infarcts), and the presence of ischemic, hemorrhagic, and vascular pathology, we used point biserial correlation coefficient. Prior to computing the correlation coefficients, we also regressed out age at death, sex, and time between DWI acquisition and death from MD and used residualized MD scores in the analysis. The p-values were adjusted for multiple comparisons using false discovery rate (FDR) correction method, with a significance threshold of p < 0.05.

#### 2.5.3 Association with cross-sectional and longitudinal cognitive performance

We then examined whether our identified latent variables from the PLS analysis were associated with ante-mortem cognitive performance. We projected the scores of each latent variable to the original data, obtaining individualized combined scores of either MD (i.e. PLS-MD) or neuropathology (i.e. PLS-neuropathology) for each participant. We then correlated these projected MD and neuropathology scores with the participants’ cognitive performance based on their Clinical Dementia Rating sum of boxes (CDR-SB) scores. Afterwards, linear mixed effects models were used to examine the relationship between the derived MD and neuropathology scores and cognitive decline trajectories for ADNI participants with longitudinal CDR-SB assessments available. In total, 114 cognitive assessments across the 21 ADNI participants were used to assess the longitudinal trajectory of cognitive deficits prior to death. We included participant ID as a random effect, and age at death, sex, and education years as covariates in the model. The interaction between individualized PLS scores and time between clinical assessment and death was used as the main variable of interest.

#### 2.5.4 Association with gene expression profiles

Finally, we examined whether the derived brain patterns corresponding to the PLS latent variables are correlated with brain wide gene expression patterns for the genes widely implicated in AD pathology (for review: [53]). We used gene expression data from the Allen Human Brain Atlas based on 6 postmortem donor brain specimens [54,55]. We then used the Brain Surrogate Maps with Autocorrelated Spatial Heterogeneity (BrainSmash; [56]) tool to calculate a null distribution for the relationship between gene expression and the LVs while accounting for spatial autocorrelation. In short, 1000 surrogate maps were computed for our LVs, and Pearson’s correlation was computed for each gene with these surrogate maps. We then calculated non-parametric p-values based on the derived null distributions, setting significance threshold to p < 0.05.

## 3. RESULTS

### 3.1 Sample characteristics

The mean time between MRI and death was 6.23 ± 2.78 years for NACC, 2.42 ± 1.6 years for ADNI, and 1.57 ± 1.0 years for AZSAND/BBDP samples. At their last evaluation, most cases were classified as dementia, with fewer having MCI or being cognitively normal. Proteinopathy and vascular pathology were present in all datasets, with adequate representation of mild to severe stages. Twenty-seven NACC cases had evidence of Lewy body pathology. The sample characteristics are presented in **Table 1**.

**Table 1.** Demographic and neuropathological characteristics of each dataset, as well as the total sample.

| Data source | NACC (97) | ADNI (21) | AZSAND (15) | Total Sample |
| --- | --- | --- | --- | --- |
| Age at death | 83.9 ± 10.4 | 81.6 ± 6.2 | 77.9 ± 8.5 | 82.9 ± 9.8 |
| Sex | 56M, 41F | 13M, 8F | 9M, 6F | 78M, 55F |
| APOE status (%positive) | 40% | 52% | 66% | 41% |
| Last visit diagnosis |  |  |  |  |
| Control | 15 | 3 | 6 | 24 |
| MCI | 14 | 5 | 2 | 21 |
| Dementia | 66 | 13 | 7 | 86 |
| Other | 2 (impaired; not MCI) |  |  |  |
| CDR-SB | 8.5 ± 6.6 | 5.9 ± 5.5 | 0.87 ± 0.9 | 7.6 ± 6.5 |
| Education | 15.5 ± 2.7 | 16.1 ± 2.7 | 15.06 ± 3.7 | 15.5 ± 2.9 |
| Thal Phase |  |  |  |  |
| 0 | 14 | 2 | 6 | 22 |
| 1 | 4 | 1 | 0 | 5 |
| 2 | 3 | 1 | 2 | 6 |
| 3 | 6 | 3 | 2 | 11 |
| 4 | 34 | 2 | 1 | 37 |
| 5 | 34 | 12 | 2 | 48 |
| Braak score |  |  |  |  |
| 0 | 1 | 0 | 0 | 1 |
| 1 | 5 | 1 | 0 | 6 |
| 2 | 15 | 4 | 1 | 20 |
| 3 | 11 | 1 | 1 | 13 |
| 4 | 15 | 3 | 6 | 24 |
| 5 | 13 | 9 | 2 | 24 |
| 6 | 35 | 3 | 3 | 41 |
| Neuritic plaque density |  |  |  |  |
| 0 | 24 | 7 | 6 | 37 |
| 1 | 15 | 2 | 0 | 17 |
| 2 | 23 | 3 | 1 | 27 |
| 3 | 33 | 9 | 6 | 48 |
| Cerebral amyloid angiopathy |  |  |  |  |
| 0 | 40 | 6 | 7 | 53 |
| 1 | 33 | 8 | 3 | 44 |
| 2 | 10 | 6 | 1 | 17 |
| 35 | 12 | 1 | 2 | 1 |
| White matter rarefaction |  |  |  |  |
| 0 | 22 | 18 | 8 | 48 |
| 1 | 37 | 2 | 1 | 40 |
| 2 | 28 | 1 | 2 | 31 |
| 3 | 8 | 0 | 2 | 10 |
| Vascular Pathology |  |  |  |  |
| Yes/No | 93/2 | 14/7 | 8/5 | 115/14 |
| Cerebral infarcts |  |  |  |  |
| Yes/No | 43/52 | 6/15 | 8/5 | 57/72 |

### 3.2 Multivariate association between mean diffusivity and neuropathology markers

Our PLS analysis revealed two significant LVs together explaining 86% of the covariance in the MD-neuropathological associations. The first LV accounted for 70% of the covariance and was significantly associated with markers of AD pathology, Thal phase (loading = 0.37, 95% CI [0.23, 0.50]), Braak score (loading = 0.33, 95% CI [0.18, 0.48]), neuritic plaque density (loading = 0.38, 95% CI [0.24, 0.51]), diffuse plaque density (loading = 0.35, 95% CI [0.21, 0.48]), and white matter rarefaction (loading = 0.18, 95% CI [0.03, 0.34]; **Figure 1)**. Regions that contributed to this LV (with p<0.05) as well as their contribution are listed in **Supplementary Table 1.** Overall, they represent medial frontal, temporal, and parietal regions that comprise the visual, frontoparietal, and salience functional networks [57].

**Figure 1.**
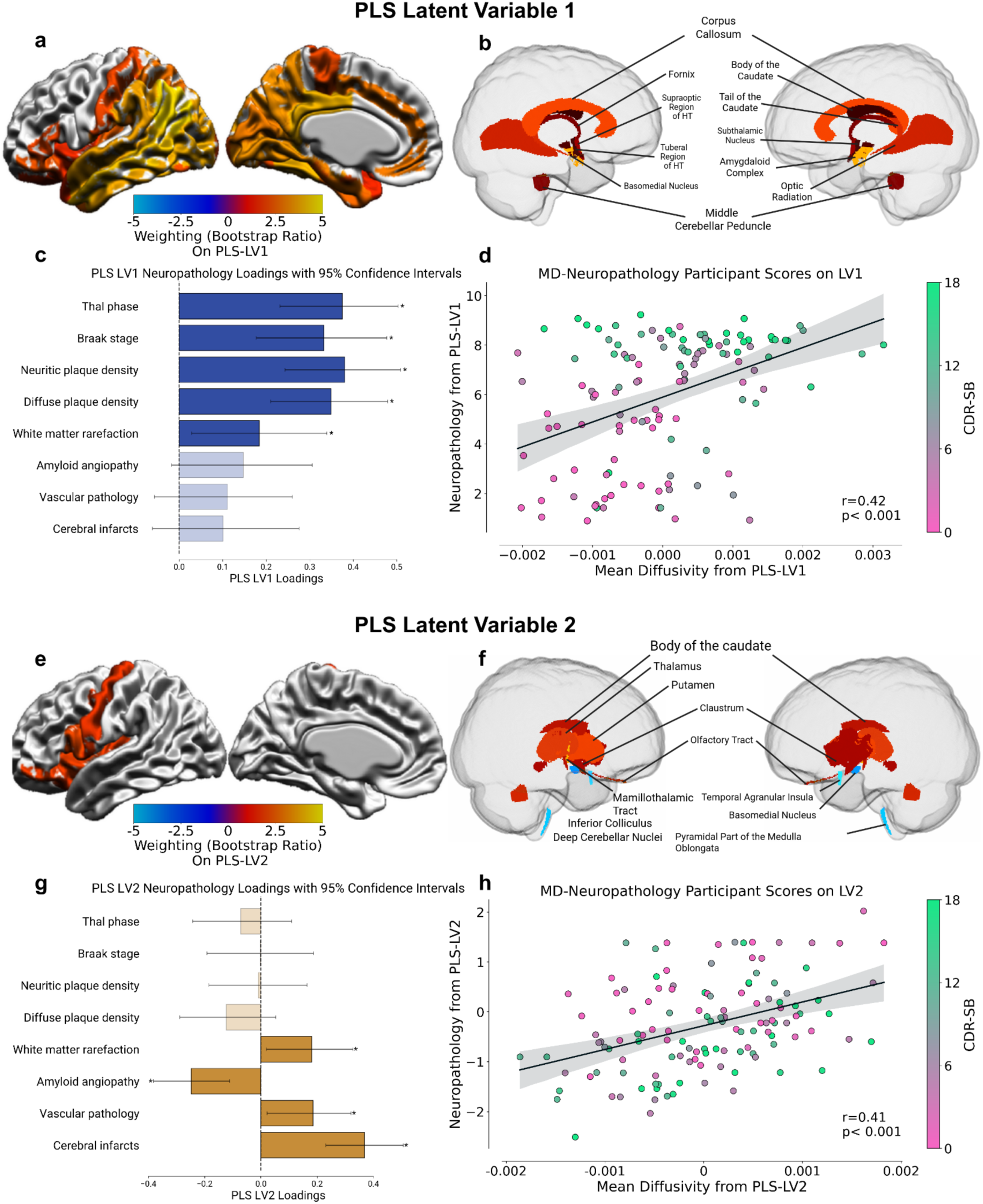
First and second latent variables (LVs) obtained from the Partial Least Squares (PLS) analysis. a-b. Mean diffusivity (MD) weight of regions that significantly contributed to LV1 (p_bootstrap_<0.05). c. Loadings LV1 neuropathological markers. d. The association between combined neuropathology and combined MD, color coded for the participants last CDR-SB score as a measure of cognition. e-f. MD weight of regions that significantly contributed to LV2 (p_bootstrap_<0.05). g. Loadings of LV2 neuropathological markers. The association between combined neuropathology and combined MD, color coded for the participants last CDR-SB score as a measure of cognition. * indicates statistical significance. CDR-SB: Clinical Dementia Rating-Sum of Boxes.

The second LV explained 16% of the covariance and was linked to vascular pathology, with significant loadings for white matter rarefaction (loading = 0.18, 95% CI [0.02, 0.33]), cerebral amyloid angiopathy (loading = -0.25, 95% CI [-0.38, -0.11]), vascular pathology (loading = 0.19, 95% CI [0.02, 0.32]), and cerebral infarcts (loading = 0.37, 95% CI [0.23, 0.51]; **Figure 1**). Regions that contributed to this latent variable (with p<0.05) as well as their contribution are listed in **Supplementary Table 2.** These regions comprise mainly the somatomotor network, as well limbic and thalamic networks [57].

### 3.3 Univariate association between mean diffusivity and neuropathology markers

When further investigating the relationship between MD and each neuropathology marker using univariate analyses. MD was significantly associated with proteinopathy biomarkers in the cortical gray matter, and with infarcts in subthalamic nuclei **(Figure 2)**. A complete list of regional correlation coefficients is included in **Supplementary Table 3**. After FDR correction, Thal Amyloid phases were associated with MD in the angular and inferior temporal gyrus, while Braak tangle score and neuritic plaque density were associated with 28 regions out of 132, mostly in the temporofrontal regions. On the other hand, the presence of any microscopic or gross infarct was significantly associated with MD in thalamic nuclei. No significant associations were found between regional MD and diffuse plaque density or cerebral white matter rarefaction.

**Figure 2.**
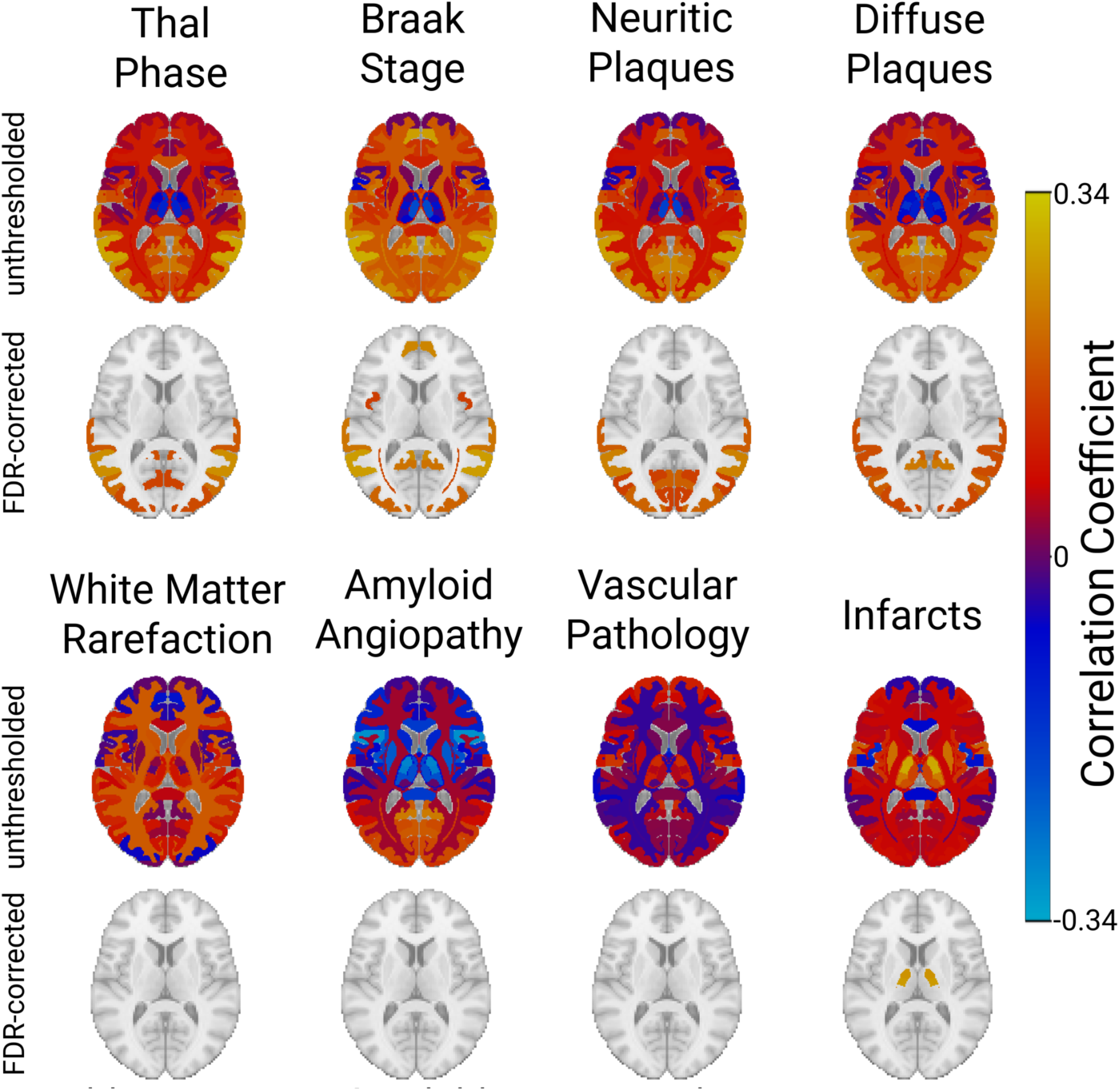
Univariate correlation maps between regional MD and neuropathology, for proteinopathies (top) and vascular pathologies (bottom). Unthresholded maps show that proteinopathies show mainly positive associations with the MD, while vascular pathologies show positive and negative correlations. Of note, only positive associations survive FDR correction.

### 3.4 Association with cognitive function

We also examined whether our identified LVs from the PLS analysis were associated with cognition. We found statistically significant associations between projected scores of MD and neuropathology for the first LV and CDR-SB (r_MD_ = 0.44, p < 0.001; r_neuropath_ = 0.59, p < 0.001; **Figure 3 - top**). Mixed effects models also showed that longitudinal changes in CDR-SB were significantly associated with MD (β = -0.28, p < 0.001) and neuropathology (β = -0.49, p < 0.001) of the identified first LV (**Figure 3 - bottom**).

**Figure 3.**
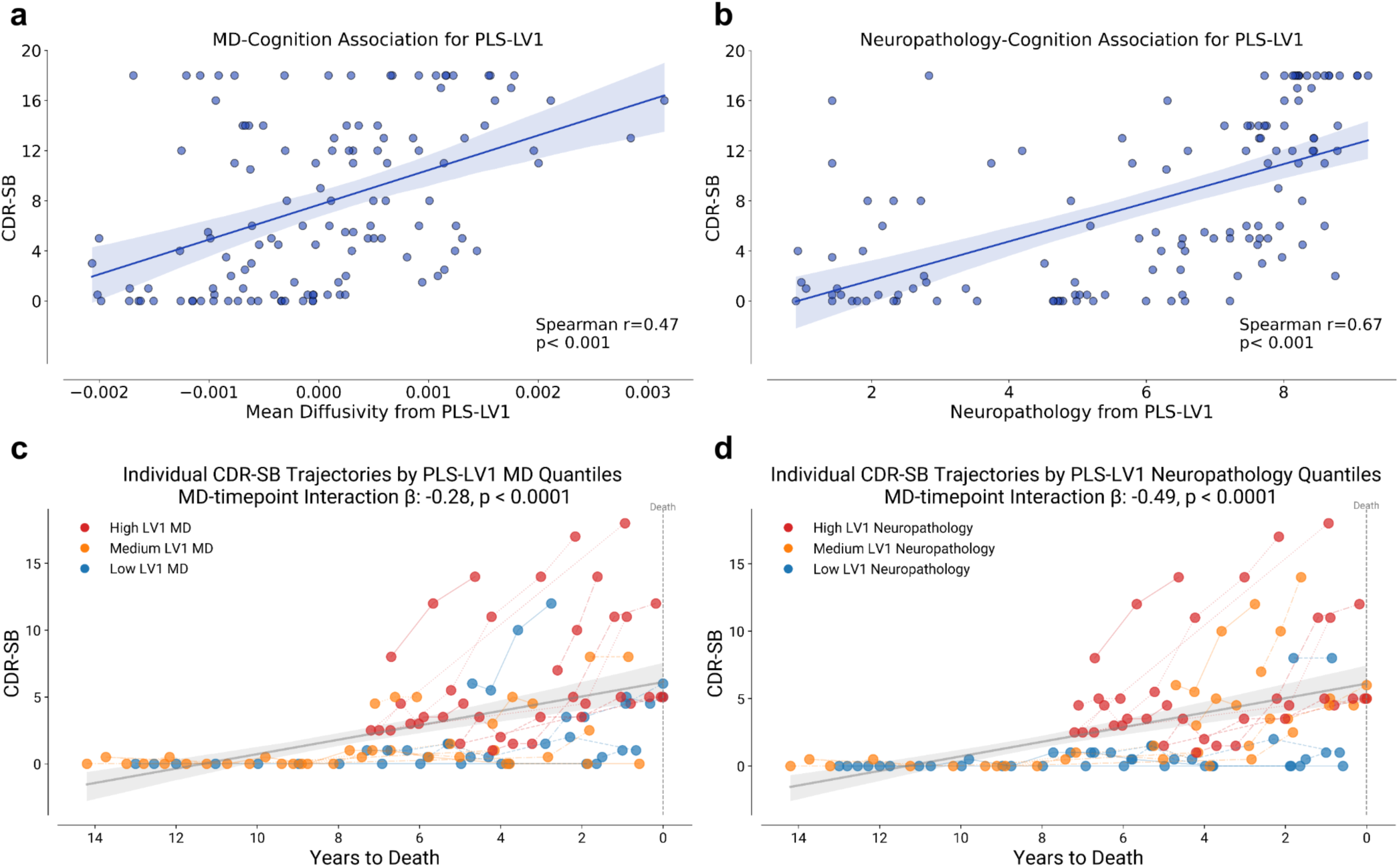
a-b Cross-sectional associations between the identified latent variable 1 (LV1) and CDR-SB at the participants’ last cognitive assessment, for MD (left) and neuropathology (right). c-d Longitudinal associations with cognitive assessments up to 14 years prior to death. CDR-SB: Clinical Dementia Rating-Sum of Boxes. PLS: Partial Least Squares.

### 3.5 Association with gene expression

MD across regions of the first LV showed significant associations with the expression of AD-related genes. Notably, MAPT, PSEN1, and TF exhibited significant positive associations with MD of the first (only for MAPT) and second LV. Interestingly, PSEN1, ADAM10, PLEKHC1, TF, and TREM2 showed negative associations with LV1 only. The genes GSK3B, CLU, ECSIT, APP, VDR, PSEN2, ABCA7, and APOE showed no statistically significant association after considering the spatial autocorrelation for correction. Correlations are presented in **Figure 4**.

**Figure 4.**
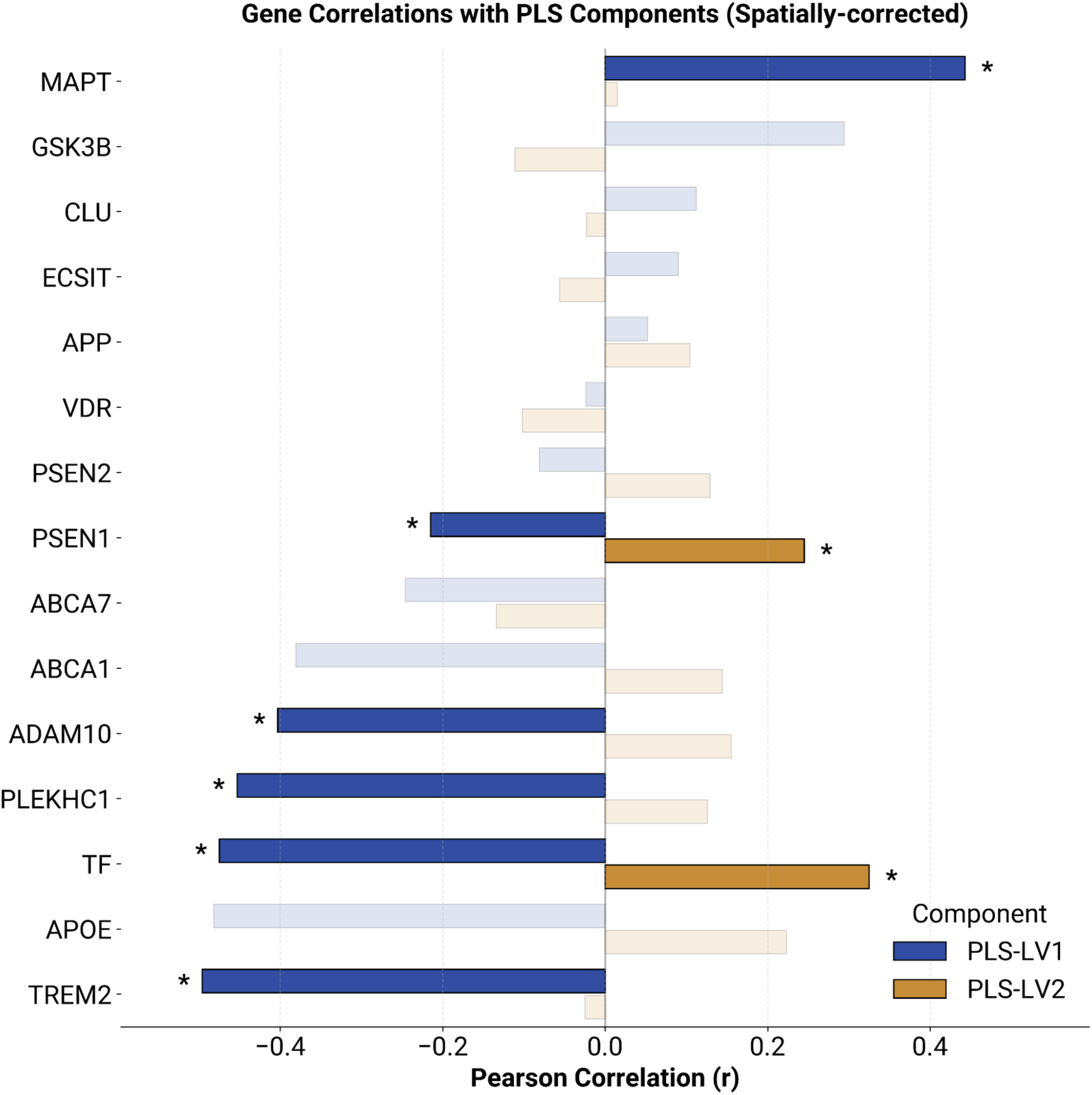
The association between regional weights on the PLS latent variables 1 and 2 and gene expression profiles, after correction for spatial autocorrelation. PLS: partial least squares, MAPT: Microtubule-Associated Protein Tau. PSEN1: Presenilin 1. TF: Transferrin. ADAM10: ADAM Metallopeptidase Domain 10. PLEKHC1: Pleckstrin Homology Domain Containing C1. TREM2: Triggering Receptor Expressed on Myeloid Cells 2. GSK3B: Glycogen Synthase Kinase 3 Beta. CLU: Clusterin. ECSIT: ECSIT Signaling Integrator. APP: Amyloid Beta Precursor Protein. VDR: Vitamin D Receptor. PSEN2: Presenilin 2. ABCA7: ATP Binding Cassette Subfamily A Member 7. APOE: Apolipoprotein E.

## 4. DISCUSSION

This clinicopathological study aimed to assess the link between MD and postmortem neuropathology markers of AD. To achieve this goal, we aggregated a large dataset from three sources (NACC, ADNI, and AZSAND/BBDP) with available cognitive assessment, in-vivo DWI, and postmortem neuropathology assessments. Our main findings demonstrate an association between DWI-derived MD and i) postmortem markers of amyloid and tau as well as vascular neuropathology and ii) cognitive status cross-sectionally at the last visit and longitudinally up to 14 years prior to death. We also observed significant associations between the neuropathology-related MD patterns and expression profiles of genes implicated in AD. Our results show a potential in validating the use of DWI as a non-invasive and more easily accessible tool for assessing the severity of AD neuropathology in combination with other emerging biomarkers.

We report an association between regional MD values and neuropathology markers for amyloid, tau, and vascular neuropathology. More specifically, our PLS analysis revealed two significant latent variables, together explaining 86% of the shared covariance between MD and neuropathology markers. Significant neuropathology markers in the first latent variable included both measures of amyloid plaque (Thal phase, CERAD neuritic and diffuse plaque score) and tau tangle (Braak stages) accumulation as well as cerebral white matter rarefaction which were associated with increased MD. Stronger associations were found in several regions of the temporal lobe and limbic system including the amygdala and hippocampus as well as in the occipital neocortex. Temporal lobe and limbic regions, in particular, may be severely impaired by both tau and amyloid pathology and these results suggest that both proteinopathies together may result in more severe microstructural damage. Additionally, the second latent variable, reflected presence of vascular pathology (e.g. ischemic or hemorrhagic) and/or any cerebral infarcts, cerebral white matter rarefaction and a negative association with cerebral amyloid angiopathy. This vascular pathology pattern was mainly associated with increased MD in the thalamus, putamen, caudate, as well as frontal and prefrontal regions. These two identified variables could be labeled as “proteinopathy” and “vascular pathology” suggesting that while MD-neuropathological associations may be mainly due to proteinopathies they are also explained by vascular changes. This pattern could be differential for markers of proteinopathies versus vascular damage in AD. Consistent with previous literature suggesting a mixed neurodegenerative and vascular etiology for white matter rarefaction in neurodegenerative diseases, and in particular AD [58], cerebral white matter rarefaction was found to contribute to both of our identified latent variables. Our univariate analyses further confirmed the associations between regional MD values and postmortem regional distribution of amyloid plaques (measured by the Thal amyloid phase) and the density of neuritic plaques as well as the distribution and density of tau pathology (measured by Braak neurofibrillary scores). These associations were mainly observed in regions of the temporal, parietal, and occipital cortex. Presence of cerebral infarcts was significantly associated with MD in the thalamus, while univariate associations with cerebral white matter rarefaction were not statistically significant.

Finally, we found strong associations between individualized PLS-MD and PLS-neuropathology scores from the first LV and CDR-SB, indicating that this variable may capture shared and unique patterns of microstructural change that lead to cognitive decline in AD. Using longitudinal cognitive assessments (CDR-SB up to 14 years prior to death), we found a significant interaction between time and PLS-MD related to changes in CDR-SB. This is consistent with previous studies showing an association between MD values and cognitive decline as well as amyloid, tau PET and CSF biomarkers associated with changes over time in these biomarkers along the AD continuum, and supports the clinical relevance of DWI-derived microstructural measures [5,12,13,15,59–62]. Therefore, MD changes might be useful for following clinical progression of AD as well as AD-type brain pathology, especially in cases with biomarker validated clinical diagnosis of AD.

Additionally, we report an association between brain-wide MD patterns from the latent variables and expression of genes known to be implicated in AD which suggests that AD-related genetic activity may influence the MD-neuropathology association reported in this study. Specifically, MAPT was positively associated with LV1, including proteinopathies, which is consistent with its role in tau pathology [63], and TF was positively associated with the LV2, including vascular markers, which is in link with the role of TF in vascular disease [64]. PSEN1, ADAM10, PLEKHC1, TF, and TREM2 showed negative associations with LV1. This is particularly interesting in the context of recent evidence showing a beneficial effect of ADAM10 in reducing amyloid beta and possibly tau pathology [65]. While the negative association with TREM2 is less clear, it has been linked to microglia and neuroinflammation which is prominent in AD [66]. Nonetheless, future studies are needed to better understand the mechanisms underlying these associations and whether these associations can be replicated based on differential gene expression in case-control studies.

Our multivariate and univariate results are in line with previous literature and suggest that the accumulation of both tau and amyloid leading to more severe neuropathology causes microstructural changes that are detectable by non-invasive in-vivo DWI biomarkers, especially in grey matter [15]. As microstructure breaks down, MD increases as a result of a lower hindrance of diffusion in a less dense environment [4,9]. Our findings are in line with previous reports by Torso and colleagues [22] that reported a relationship between gray matter microstructural changes and both amyloid and tau pathology mainly involving temporal and limbic regions in 43 autopsy cases. We have extended this work in a larger sample size and specifically highlighted the contribution of both proteinopathies and vascular changes to MD values. Associations were found in regions involved in both early and advanced disease stages suggesting that early amyloid deposition and neuroinflammatory changes as well as later neuronal and synaptic dysruption may lead to detectable changes in the observed MD. Previous work demonstrated an association between DWI measures and cortical architecture and altered cellular organisation, in particular with mini-columnar thinning which has been associated with aging and dementia [22,67,68].

We acknowledge some limitations of this work. First, the time between the last MRI session and death/autopsy (mean time of 5 years) may influence the results drawn from imaging as neuropathological changes will continue to progress during this period. However, we accounted for this time difference by either including time between MRI and death as a covariate, or regressing it out from the MD values. We further repeated the analyses in the subset of the cases with time between MRI and death less than five years, obtaining very similar results (data not shown). While we used data collected from several sites, both imaging protocols and neuropathology assessments were similar between data sources and we applied the same processing steps on all in-vivo DWI scans and harmonized them using the widely used neuroCombat framework while preserving effects of interest (i.e. variability across subjects, brain regions, and biological sex), allowing us to evaluate a relatively large cohort. Further, this is a cross-sectional study, and future studies should examine the longitudinal change in MD and its link with neuropathology in AD and other neurodegenerative diseases. Lastly, our gene expression profiles come from healthy specimens with no history of clinical symptoms. Future work should also attempt to examine expression levels in the same specimens for which neuropathology assessments are available, to better explain MD-neuropathology-genetic associations. Nevertheless, clinicopathological correlation studies comparing MD values to neuropathology assessments, which remain the gold standard for the diagnosis of neurodegenerative diseases, are sparse and critically needed to better understand its clinical utility.

In conclusion, this clinicopathological study provides evidence for a link between in vivo MD in cortical and subcortical gray matter regions and postmortem neuropathology markers of AD. By aggregating and harmonizing data from NACC, ADNI, and AZSAND/BBDP, we demonstrated that MD is associated not only with amyloid, tau, and vascular pathology, but also with cognitive status both at the final clinical assessment and retrospectively up to 14 years before death. Finally, the observed MD patterns related to neuropathology were linked to gene expression profiles implicated in AD. These findings support the potential of DWI-derived MD as a non-invasive and accessible biomarker for assessing AD-related brain pathology.

## Data Availability

All data produced are available online at https://naccdata.org/ and https://adni.loni.usc.edu/. Please refer to ADRC contributing sites for more information when needed.

## 6. CONSENT STATEMENT

Written informed consent was obtained from all participants in accordance with protocols approved by the respective local Institutional Review Boards (IRBs). For participants enrolled through the National Institute on Aging Alzheimer’s Disease Centers (ADRCs), consent was administered and documented under the oversight of each center’s IRB prior to data submission to the National Alzheimer’s Coordinating Center (NACC). Similarly, participants from the Alzheimer’s Disease Neuroimaging Initiative (ADNI) and the Arizona Study of Aging and Neurodegenerative Disorders (AZSAND) provided written informed consent under IRB-approved protocols specific to each study site. All procedures adhered to the ethical principles outlined in the Declaration of Helsinki and relevant regulatory guidelines governing human subjects research.

## 7. ACKNOWLEDGEMENTS

Data used in this study were provided from three sources including NACC, ADNI and AZSAND/BBDP. The authors would like to thank all the volunteer donors who participated in these studies and donated their brains, as well as their families. We also thank the personnel and researchers who helped contribute clinical data including MRI scans and postmortem brains from the participants in these three programs.

We express appreciation to the contributors of NACC. The NACC database is funded by NIA/NIH Grant U24 AG072122. Standardized Centralized Alzheimer’s & Related Dementias Neuroimaging (SCAN) is a multi-institutional project funded as a U24 grant (AG067418) by the National Institute on Aging in May 2020. Data collected by SCAN and shared by NACC are contributed by the NIA-funded ADRCs and affiliated centers as follows: Arizona Alzheimer’s Center – P30 AG072980, R01 AG069453, P30 AG019610 (PI: Eric Reiman, MD), and the State of Arizona; Boston University – P30 AG013846 (PI: Neil Kowall, MD); Cleveland ADRC – P30 AG062428 (PI: James Leverenz, MD); Cleveland Clinic, Las Vegas – P20 AG068053; Columbia – P50 AG008702 (PI: Scott Small, MD); Duke/UNC ADRC – P30 AG072958 (PI: Heather Whitson, MD); Emory University – P30 AG066511 (PI: Allan Levey, MD, PhD); Indiana University – R01 AG19771, P30 AG10133, P30 AG072976 (PI: Andrew Saykin, PsyD), R01 AG061788 (PI: Shannon Risacher, PhD), R01 AG053993 (PI: Yu-Chien Wu, MD, PhD), U01 AG057195 (PI: Liana Apostolova, MD), U19 AG063911 (PI: Bradley Boeve, MD), and support from the Department of Radiology and Imaging Sciences; Johns Hopkins – P30 AG066507 (PI: Marilyn Albert, PhD); Mayo Clinic – P50 AG016574 (PI: Ronald Petersen, MD, PhD); Mount Sinai – P30 AG066514 (PI: Mary Sano, PhD), R01 AG054110 and R01 AG053509 (PI: Trey Hedden, PhD); New York University – P30 AG066512-01S2 (PI: Thomas Wisniewski, MD), R01 AG056031 (PI: Ricardo Osorio, MD), R01 AG056531 (PIs: Ricardo Osorio, MD; Girardin Jean-Louis, PhD); Northwestern University – P30 AG013854 (PI: Robert Vassar, PhD), R01 AG045571, R56 AG045571, R01 AG067781, U19 AG073153, R01 AG077444, R01 AG056258 (PI: Emily Rogalski, PhD), R01 DC008552 (PI: M.-Marsel Mesulam, MD), R01 NS075075 (PI: Emily Rogalski, PhD); Oregon Health and Science University – P30 AG008017, R56 AG074321 (PI: Jeffrey Kaye, MD); Rush University – P30 AG010161 (PI: David Bennett, MD); Stanford – P30 AG066515, P50 AG047366 (PI: Victor Henderson, MD, MS); University of Alabama, Birmingham – P20 AG086401 (PI: Erik Roberson, MD, PhD); University of California, Davis – P30 AG10129, P30 AG072972 (PI: Charles DeCarli, MD); University of California, Irvine – P50 AG016573, P30 AG066519 (PI: Frank LaFerla, PhD); University of California, San Diego – P30 AG062429 (PI: James Brewer, MD, PhD); University of California, San Francisco – P30 AG062422 (PI: Gil Rabinovici, MD); University of Kansas – P30 AG035982 (PI: Russell Swerdlow, MD); University of Kentucky – P30 AG028283-15S1, P30 AG072946 (PI: Linda Van Eldik, PhD; Brian Gold, PhD); University of Michigan – P30 AG053760, P30 AG072931 (PI: Henry Paulson, MD, PhD), Cure Alzheimer’s Fund 200775, U19 NS120384 (Site PI: Henry Paulson, MD, PhD), R01 AG068338 (MPIs: Bruno Giordani, PhD; Carol Persad, PhD; Yi Murphey, PhD), S10 OD026738-01 (PI: Douglas Noll, PhD), R01 AG058724, R35 AG072262, W81XWH2110743, R01 AG073235, 1I01 RX001534, IRX001381 (PI: Benjamin Hampstead, PhD); University of New Mexico – P20 AG068077, P30 AG086404 (PI: Gary Rosenberg, MD); University of Pennsylvania – P30 AG072979, State of PA project 2019NF4100087335, Rooney Family Research Fund, R01 AG055005 (PI: David Wolk, MD); University of Pittsburgh – P50 AG005133, P30 AG066468 (PI: Oscar Lopez, MD); University of Southern California – P50 AG005142, P30 AG066530 (PI: Helena Chui, MD); University of Washington – P50 AG005136, P30 AG066509 (PI: Thomas Grabowski, MD); University of Wisconsin – P50 AG033514, P30 AG062715 (PI: Sanjay Asthana, MD, FRCP); Vanderbilt University – P20 AG068082 (PI: Angela Jefferson, PhD); Wake Forest – P30 AG072947 (PI: Suzanne Craft, PhD); Washington University, St. Louis – P01 AG003991, P01 AG026276, P30 AG066444 (PI: John Morris, MD), R01 AG021910 (PI: Randy Buckner), R01 AG043434 (PI: Catherine Roe), P20 MH071616, P30 NS098577, R01 EB009352 (PI: Dan Marcus), UL1 TR000448 (PI: Brad Evanoff); Yale – P50 AG047270, P30 AG066508 (PI: Stephen Strittmatter, MD, PhD), R01 AG052560 (MPIs: Christopher van Dyck, MD; Richard Carson, PhD), R01 AG062276 (PI: Christopher van Dyck, MD); and 1 Florida – P30 AG066506-03 (PI: Glenn Smith, PhD), P50 AG047266 (PI: Todd Golde, MD, PhD).

We also express appreciation to contributors of ADNI. Data collection and sharing for this project was funded by the Alzheimer’s Disease Neuroimaging Initiative (ADNI) (National Institutes of Health Grant U01 AG024904) and DOD ADNI (Department of Defense award number W81XWH-12-2-0012). ADNI is funded by the National Institute on Aging, the National Institute of Biomedical Imaging and Bioengineering, and through generous contributions from the following: AbbVie, Alzheimer’s Association; Alzheimer’s Drug Discovery Foundation; Araclon Biotech; BioClinica, Inc.; Biogen; Bristol-Myers Squibb Company; CereSpir, Inc.; Cogstate; Eisai Inc.; Elan Pharmaceuticals, Inc.; Eli Lilly and Company; EuroImmun; F. Hoffmann-La Roche Ltd and its affiliated company Genentech, Inc.; Fujirebio; GE Healthcare; IXICO Ltd.; Janssen Alzheimer Immunotherapy Research & Development, LLC.; Johnson & Johnson Pharmaceutical Research & Development LLC.; Lumosity; Lundbeck; Merck & Co., Inc.; Meso Scale Diagnostics, LLC.; NeuroRx Research; Neurotrack Technologies; Novartis Pharmaceuticals Corporation; Pfizer Inc.; Piramal Imaging; Servier; Takeda Pharmaceutical Company; and Transition Therapeutics. The Canadian Institutes of Health Research is providing funds to support ADNI clinical sites in Canada. Private sector contributions are facilitated by the Foundation for the National Institutes of Health (www.fnih.org). The grantee organization is the Northern California Institute for Research and Education, and the study is coordinated by the Alzheimer’s Therapeutic Research Institute at the University of Southern California. ADNI data are disseminated by the Laboratory for NeuroImaging at the University of Southern California.

We additionally thank the Arizona Study of Aging and Neurodegenerative Disorders and Brain and Body Donation Program (AZSAND/BBDP) that has been supported by the National Institute of Neurological Disorders and Stroke (U24 NS072026 National Brain and Tissue Resource for Parkinson’s Disease and Related Disorders), the National Institute on Aging (P30 AG19610 and P30AG072980, Arizona Alzheimer’s Disease Center), the Arizona Department of Health Services (contract 211002, Arizona Alzheimer’s Research Center), the Arizona Biomedical Research Commission (contracts 4001, 0011, 05-901, and 1001 to the Arizona Parkinson’s Disease Consortium), and the Michael J. Fox Foundation for Parkinson’s Research.

## 8. CONFLICTS OF INTERESTS

The authors declare no conflict of interest.

## 9. FUNDING SOURCES

ZA is supported by Brain Canada through its Rising Star program as well as the Natural Sciences and Engineering Research Council of Canada (NSERC) Vanier Scholars program. CT has been supported by a postdoctoral fellowship from the Canadian Institutes of Health Research (CIHR). RM is supported by a scholarship from FRQS. MD reports receiving research funding from the FRQS (https://doi.org/10.69777/330750), NSERC, CIHR, and Brain Canada. YZ reports receiving research funding from the FRQS (https://doi.org/10.69777/320107), NSERC, and CIHR. The authors also acknowledge use of Compute Canada (https://alliancecan.ca/en) resources for performing the image processing analyses in the presented work.

## Notes

### Competing Interest Statement

The authors have declared no competing interest.

### Author Declarations

We aggregated participants' data from the NACC (September 2024 data freeze; https://naccdata.org/), ADNI (https://adni.loni.usc.edu/) a longitudinal multicenter observational study aiming to validate biomarkers of AD, as well as additional participants from the Arizona Study of Aging and Neurodegenerative Disorders (AZSAND) and its Brain and Body Donation Program (BBDP), a longitudinal clinicopathological study at Banner Sun Health Research Institute (BSHRI) (http://www.brainandbodydonationprogram.org/). The AZSAND/BBDP dataset has been de-identified at the source, and we worked with de-identified data.

